# Distinct Gastrointestinal Symptom Phenotypes in Adults with Cystic Fibrosis are linked to complications and medication use

**DOI:** 10.64898/2026.01.16.26344251

**Authors:** Jemila R Holaman, Darren J Sills, Hisham A Saumtally, Charlotte C Johnson, Arantxa A Recto, Ryan Marsh, Andrew Prayle, Tanya M Monaghan, Luca Marciani, Robin C Spiller, Helen I Barr, Damian G Downey, Christopher van der Gast, Daniel Peckham, Iain Stewart, Alan R Smyth

## Abstract

**Background and aims:** Gastrointestinal symptoms remain common in adults with cystic fibrosis (CF) despite cystic fibrosis transmembrane conductance regulator modulator use, suggesting persistent and heterogeneous gut dysfunction. This prospective observational study tests the hypothesis that distinct gut symptom phenotypes in CF can be observed and linked to underlying mechanisms.

**Methods:** Adults from three UK CF centres completed the Gastrointestinal Symptom Rating Scale, Patient Assessment of Constipation Symptoms and a bowel-habit questionnaire. Latent class analysis using an ordinal logistic model was applied to 36 symptom indicators. Associations between phenotypes and demographic, clinical, and treatment variables were examined using generalised linear models.

**Results:** Three hundred participants completed questionnaires (54% male; median 31 years). We identified four symptom phenotypes: mild; moderate-constipation predominant; moderate-diarrhoea predominant and severe. The severe phenotype was associated with gastroesophageal reflux (RRR 2.86; 95%CI: 1.30-6.31; p=0.009), distal intestinal obstruction syndrome (RRR 2.46; 95%CI: 1.04-5.81; p=0.04), proton pump inhibitor (RRR 3.29; 95%CI 1.39-7.74; p=0.007), and laxative use (RRR 6.13; 95%CI 2.54-14.84; p<0.001). CF-related liver disease was associated with both moderate-constipation and diarrhoea phenotypes, respectively (RRR 2.08; 95%CI 1.13-3.81; p=0.018; RRR 2.11; 95%CI 1.03-4.29; p=0.04). There was a lower likelihood of long-term oral antibiotic use in the moderate-constipation phenotype (RRR 0.53; 95%CI 0.3-0.92; p=0.025) and moderate-diarrhoea phenotype (RRR 0.46; 95%CI 0.24-0.91; p=0.025).

**Conclusions:** Four distinct symptom phenotypes were identified, independent of demographics and pancreatic status, but associated with specific complications and medication profiles. These phenotypes provide a framework for mechanistic studies within the GRAMPUS-CF cohort and precision management of CF-related gut disease.

## INTRODUCTION

Cystic fibrosis transmembrane conductance regulator (CFTR) modulator therapies have markedly improved pulmonary outcomes, including enhanced lung function and reduced infection rates (1). However, many people living with cystic fibrosis (PwCF) continue to experience gastrointestinal (GI) symptoms, including bloating, pain, wind and loose stools, that can persist despite these therapies (2–4). The manifestation of GI symptoms varies considerably among individuals, and the CF community has consistently highlighted the need for research into effective treatments to be a high priority in recent international surveys (3, 5, 6). Despite this, the pathophysiology of these symptoms and complications in CF remains poorly understood, and outcome measures used in clinical studies may inadequately capture GI symptoms (3, 7), limiting the ability to evaluate therapeutic response.

GI physiology in PwCF is characterised by multiple abnormalities that may contribute to symptom burden, including pancreatic insufficiency, reduced bicarbonate secretion into the duodenum (8), abnormal mucus structure, gut dysbiosis, intestinal inflammation (9) and altered motility (10). Together, these mechanisms create a multifactorial physiological environment in which symptoms often overlap rather than occur in isolation.

A symptom cluster is defined as two or more related symptoms that manifest simultaneously (11, 12). Investigating distinct GI symptom clusters in PwCF, and the pathogenic mechanisms associated within GI phenotypes, offers a precision-medicine approach to directly target relevant GI physiology and reduce symptom burden. This phenotyping has proven valuable in other chronic GI disorders (13–15) and may reveal mechanistically distinct subgroups within the CF population that could benefit from tailored therapeutic strategies.

PwCF have, until the introduction of CFTR modulator therapies been advised and habituated to consume an unrestricted high-calorie, high-fat diet, often called the “legacy CF diet” (16). Additionally, the “legacy CF diet” is characterised by poor diet quality (16); which may accentuate GI symptoms, dysbiosis and intestinal inflammation (17–19).

Patient-reported outcome measures have been developed to capture GI symptom severity; however, symptoms rarely occur in isolation. Reliance on unidimensional scoring systems may not fully capture the complexity of co-occurring symptoms, necessitating the use of individualised approaches using Latent Class Analysis (LCA) to identify symptom phenotypes. Recent studies using LCA identified irritable bowel syndrome phenotypes, offering novel insights into underlying mechanisms and informing personalised treatment approaches (13–15). Despite the high prevalence and clinical importance of GI symptoms in CF, no studies have used multivariate clustering methods to systematically define symptom phenotypes or investigate their mechanistic basis.

Gut Research Advancing a Mechanistic and Personalised Understanding of Symptoms in Cystic Fibrosis (GRAMPUS-CF) is a prospective, longitudinal observational study to determine phenotypes of GI symptoms based on co-occurring GI symptoms in PwCF and elucidate underlying mechanisms. Specifically, we hypothesised that LCA would identify distinct, clinically meaningful GI symptom phenotypes that are associated with specific clinical characteristics, complications and therapeutic requirements. Here we report the study design, baseline characteristics, multivariate classification of symptoms and phenotypic features using LCA, and phenotypic features of the identified symptom classes.

## METHODS

### Study design

The participants described here were enrolled into the main cohort (Group A). Group A participants could also enrol into one of two nested GRAMPUS-CF sub-groups (group B and C) depending on their preference, with progressively detailed data collection (supplementary Figure S1). Group A participants completed three symptom questionnaires and 24-hour dietary recall only. These data were collected on three occasions, six months apart. Data collection allowed a three-month window to account for clinic scheduling variability. This pre-specified window accommodated clinic visits while minimising potential bias in the longitudinal data. Group B participants, a subgroup of Group A, also provided blood and stool samples for inflammatory and microbiome analyses. Group C participants, a further subgroup of group B, underwent a magnetic resonance imaging (MRI) scanning protocol on a single study day. Data from stool microbiome, MRI and blood inflammatory markers in will be presented in future publications.

This nested design allowed participants to engage at a level matching their capacity and interest, whilst enabling comprehensive mechanistic investigation in a subset. Once we identified phenotypes of GI symptoms, we tested whether these phenotypes were associated with an increased prevalence of GI complications such as: CF-related liver disease, gastroesophageal reflux disease, distal intestinal obstruction syndrome, meconium ileus, CF diabetes and pancreatic exocrine insufficiency. Furthermore, we planned to determine whether GI phenotypes were associated with the prescription of concurrent medicines such as: CFTR modulators, oral antibiotics, immunosuppressants, proton pump inhibitors, laxatives and nutritional supplements.

### Participants

Participants were enrolled from one of three specialist CF centres in the United Kingdom: Nottingham University Hospitals NHS Trust, Leeds Teaching Hospitals NHS Trust and Belfast Health and Social Care Trust. All individuals attending a clinical visit were approached consecutively, regardless of their symptom history, unless the clinical team deemed a patient’s participation inappropriate, such as acute illness at that time. After providing detailed study information, written informed consent was requested. The participant could consent to participation in Group A (questionnaire), group B (Group A plus blood and stool samples) or group C (sub-group B plus a day of MRI scanning) based on individual preference. The nested subgroups are shown in supplementary Figure S1. Eligible participants were aged 16 years or over with a confirmed CF diagnosis. Patients were excluded if they had an established pre-existing gastrointestinal condition (e.g. inflammatory bowel disease, coeliac disease or gastrointestinal cancer) or were pregnant. Recruitment commenced on the 25^th^ October 2023 and was completed on the 4^th^ April 2025.

Baseline sampling has been completed, LCA and clinical characteristics analysis is presented herein this paper. Additional gut microbiome, serum inflammatory markers and MRI data will be presented in subsequent papers.

### Data collection

#### Demographic and clinical data

These data were collected from the medical records and from the UK CF Registry database. Data were collated and managed using REDCap (20, 21). Collected variables included age, sex, ethnicity, CF genotype (categorised as F508del homozygous, F508del heterozygous, or other), CF related co-morbidities, pancreatic exocrine insufficiency status, relevant gastrointestinal medical history, respiratory colonisation, pulmonary function; Forced Expiratory Volume in one second (FEV_₁_) percentage predicted and Forced Vital Capacity (FVC) percentage predicted, anthropometric measures and concurrent medications.

To protect participant confidentiality in the context of a small sample, certain potentially identifying variables were not reported. All analyses were used the full dataset, but only non-identifiable aggregate results, where n> 5 are presented.

#### Participant questionnaires

Participants completed symptom questionnaires either electronically or on paper, during a baseline face-to-face visit. Additionally, those enrolled at selected recruiting sites could complete questionnaires via a smartphone application. Questionnaire data were captured in REDCap. Symptom assessment measures included validated questionnaires: the Patient Assessment of Constipation Symptoms (22, 23) (PAC-SYM) and the Gastrointestinal Symptom Rating Scale (24, 25) (GSRS) (supplemental Table S1). Seven supplementary questions (GRAMPUS-7), including the Bristol Stool Form Scale (BSFS) (26), were added to address symptoms relating to bowel habit not covered by the validated questionnaires (supplemental methods, Table S1). All questionnaires used a seven day recall period.

#### Dietary recall

Participants recorded all food and drinks consumed on the Intake24 (27) platform (intake24.org, UK, V3 2023). Intake24 is an open-source web-based dietary assessment research tool based on the 24-hour recall method, designed for self-completion. Intake24 is a validated platform to capture dietary intake, containing branded and unbranded food products, household measures and pictorial portion sizes. Participants were asked to complete Intake24 at least once, and ideally twice, at each timepoint. Recalls were not required to be consecutive but were intended to capture intake within the defined timepoint window.

### Ethical approval

The study was prospectively registered on clinicaltrials.gov (NCT05934656), prior to commencing enrolment, and approved by the Cambridge South Research Ethics Committee (23/EE/0092) on the 14^th^ June 2023. The study was carried out according to the principles of Good Clinical Practice and in accordance with the Declaration of Helsinki. Participants provided fully informed written consent prior to undertaking any study related procedure. Local regulatory approvals were obtained at each participating site.

### Statistical methods

#### Sample size

The primary outcome was identification of distinct GI symptom phenotypes using LCA. The minimum sample size for Group A was 300, selected pragmatically based on simulation studies in the literature to differentiate at least two clinically meaningful classes in multivariate analysis (28). In sub-group B, a target sample size of 100 was estimated to provide 80% power to detect a 50% relative difference in abundance in the top 10 microbial taxa, assuming two equally sized classes of 50. Sub-group C, the planned recruitment target was 36 participants, providing 80% power to detect a difference in small bowel water content area under the curve, assuming two classes of 18, with our prior study Gut Imaging for Function & Transit in CF (10) demonstrating a standardised effect size (Cohen’s D) of 0.97 between the healthy and the CF gut (29).

#### Dietary data

Dietary recall data were exported from Intake24 and nutrient intakes calculated using the UK Nutrient Databank. Initial data cleaning involved identifying missing food items and assessing plausibility of reported energy intakes and portion sizes. Nutritional supplements, including protein/energy products and vitamin or mineral preparations, were excluded from the primary dietary analysis. Data quality and completeness checks were performed in RStudio in accordance with the Intake24 Data Cleaning Guidelines (Version 1.0, 2023). Additional details are provided in the supplementary materials. Daily average nutritional intake was calculated for each participant.

Two dietary pattern indices (30, 31) were derived from Intake24 data. The Dietary inflammatory index (DII) estimates the overall inflammatory potential of the diet by scoring nutrients and food components according to their associations with inflammatory biomarkers (30, 32, 33). In GRAMPUS-CF, Intake24 data provided data for 27 of the 45 possible DII components. A higher DII score is indicative of a more pro-inflammatory diet (32). The use of 27 of the 45 nutrients gives a DII score which ranges from −5.5 to +5.5 (30). The diet quality index – international (DQI-I) was developed to assess overall dietary quality across four domains: variety, adequacy, moderation and balance (31). Each dimension contributes to a total score ranging from 0 to 100, with higher values indicating better diet quality. Both DII and DQI-I scores were calculated in RStudio, following established scoring procedures (31, 32).

#### Latent class analysis

LCA was performed using responses to items from all GI symptom questionnaires. LCA is a probabilistic, model-based form of unsupervised clustering, that identifies unobserved (latent) subgroups based on patterns of response probabilities across indicator variables (28). The probability of class membership for each participant is calculated; class was assigned according to highest probability (34). To support model convergence and ensure comparability across measures, all symptom items were standardised to a 3-point Likert unidirectional scale of Mild (1), Moderate (2) and Severe (3) (supplemental methods). Bidirectional questions including frequency of bowel movement and stool softness chart were recoded as two indicators, respectively (see supplemental methods, Table S2). A total of 36 indicators based on 34 items were included.

Missing indicator responses were recoded as Mild (1), the median response and assuming absence of severe symptoms when data were not provided.

Models with 1 to 6 latent classes were fitted and compared using the Polytomous Variable Latent Analysis package in R version 4.3.0, specified with a logistic ordinal model. Model fit and the optimal model was selected based on: (i) low Akaike Information Criterion (AIC) and Bayesian Information Criterion (BIC) values, (ii) balanced high entropy indicating clear class separation with minimised overfitting, and each class representing at least 10% of the cohort to ensure class stability, and support clinical relevance through easily recognisable symptom patterns. Indicator posterior probabilities on the class were used to label the phenotype.

#### Statistical comparisons

Data were described using frequency and proportion, median and interquartile range (IQR) for non-normal data, or mean and standard deviation (SD) for normal continuous data. ANOVA, Chi-square and Kruskal-Wallis were used to interpret differences across phenotypes, using post hoc tests for between group differences, two-sided significance level of α = 0.05 was applied throughout, p<0.05 was defined as statistical significance. Associations between phenotype and clinical characteristics were tested using multinomial logistic regression, estimating the relative risk ratio (RRR) compared with a base group, reported with 95% confidence intervals (CI) from robust standard errors derived with variance sandwich estimators.

## RESULTS

### Study cohort

Three hundred adults with CF completed baseline questionnaires (Group A), full details in Figure 1. There were 162 males and 138 females with a median age of 31 years (IQR 24 to 40) (Table 1). Most participants (95.7%, n=287) were of white ethnicity and had at least one copy of the F508del variant (92.3%, n=277). Most participants had pancreatic exocrine insufficiency (88.3%, n=265) and were prescribed pancreatic enzyme replacement therapy (84%, n=252). Sweat chloride levels were available for 64.7% of participants (n = 194), with a median value of 47.1 mmol/L (IQR 32 to 76.2), reflecting a mix of diagnostic and therapy monitoring values. A sweat chloride of ≥ 60 mmol/L is diagnostic of CF, but values drop considerably in response to CFTR modulators. . CFTR modulator therapy was prescribed in 84.3% participants (n = 253), with triple modulator therapy (elexacaftor/tezacaftor/ivacaftor) being the most common regimen (80.3%, n=241). Concurrent medications at baseline are shown in (Tables 1 and 2).

**Figure 1.**
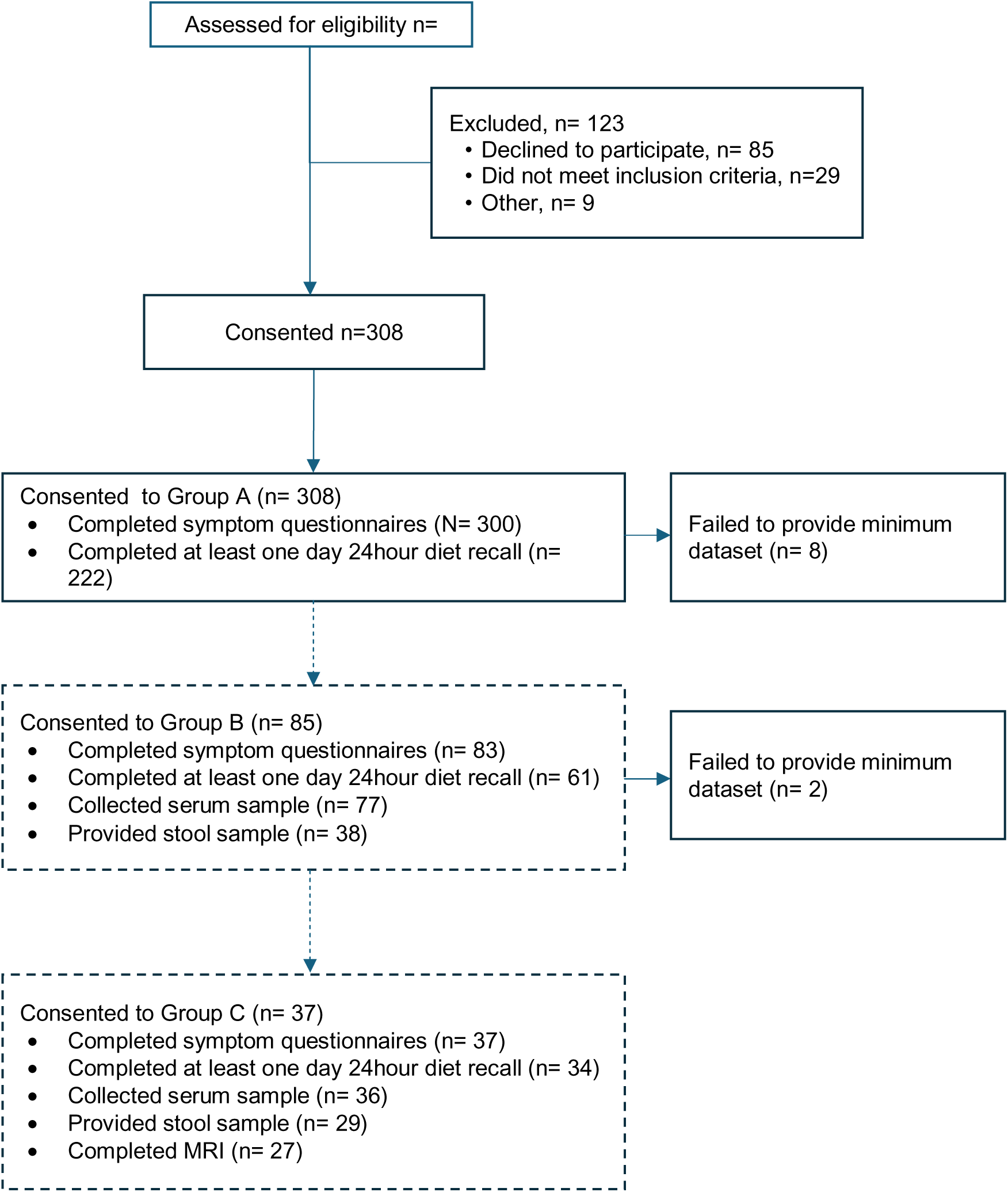
Participant enrolment, group allocation and flow of questionnaire completion. This diagram outlines the number of individuals screened, reasons for exclusion, allocation into study groups and completion of symptom questionnaires across study groups. Dashed lines represent tiered nested sub-groups

**Table 1 -.**
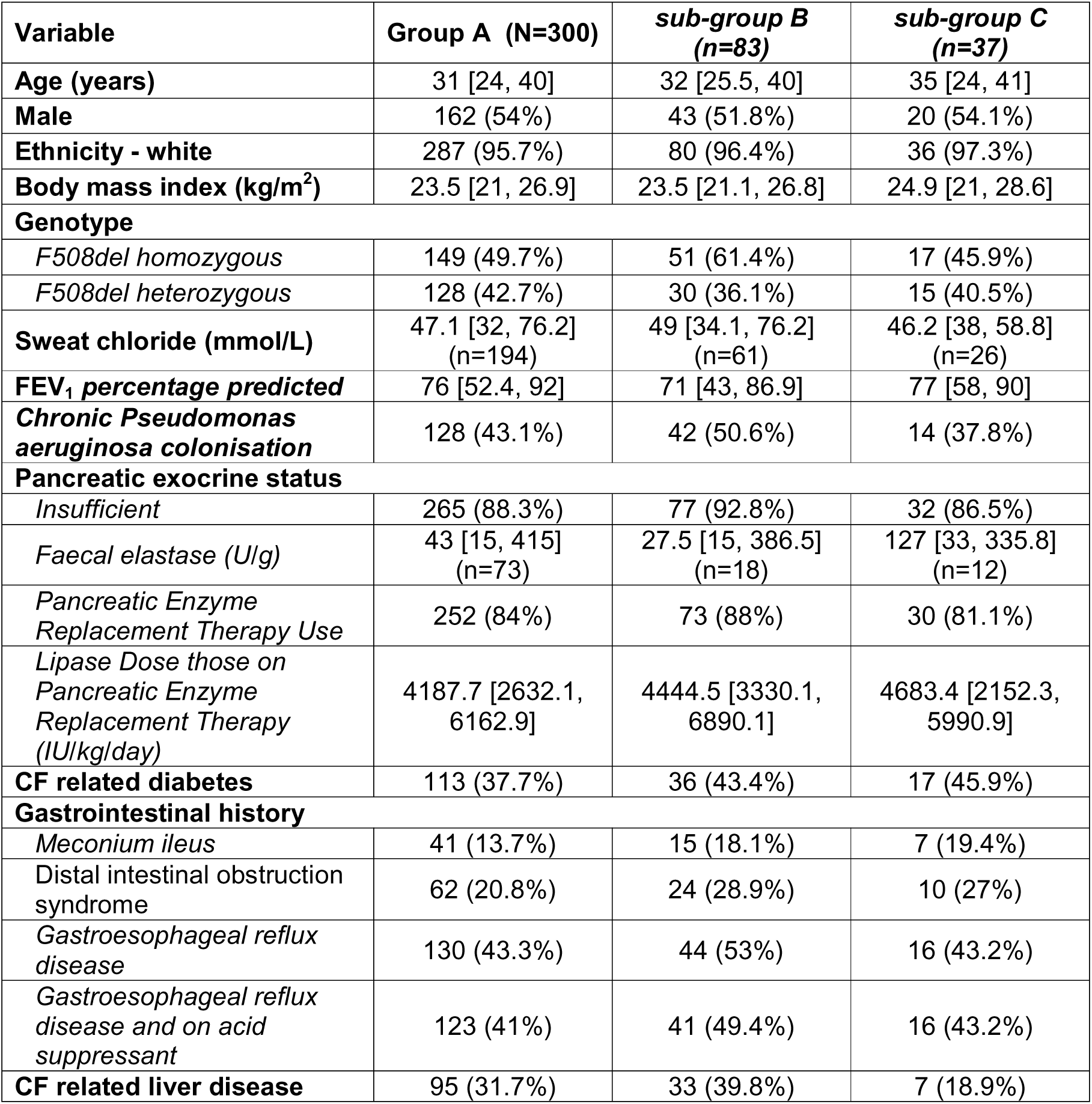
Baseline demographic and clinical characteristics stratified by study group. Results are described as mean (SD), median [interquartile range] or number (%).

| Variable | Group A (N=300) | sub-group B (n=83) | sub-group C (n=37) |
| --- | --- | --- | --- |
| **Age (years)** | 31 [24, 40] | 32 [25.5, 40] | 35 [24, 41] |
| **Male** | 162 (54%) | 43 (51.8%) | 20 (54.1%) |
| **Ethnicity - white** | 287 (95.7%) | 80 (96.4%) | 36 (97.3%) |
| **Body mass index (kg/m<sup>2</sup>)** | 23.5 [21, 26.9] | 23.5 [21.1, 26.8] | 24.9 [21, 28.6] |
| **Genotype** |  |  |  |
| *F508del homozygous* | 149 (49.7%) | 51 (61.4%) | 17 (45.9%) |
| *F508del heterozygous* | 128 (42.7%) | 30 (36.1%) | 15 (40.5%) |
| **Sweat chloride (mmol/L)** | 47.1 [32, 76.2]<br>(n=194) | 49 [34.1, 76.2]<br>(n=61) | 46.2 [38, 58.8]<br>(n=26) |
| **FEV<sub>1</sub> percentage predicted** | 76 [52.4, 92] | 71 [43, 86.9] | 77 [58, 90] |
| **Chronic *Pseudomonas aeruginosa* colonisation** | 128 (43.1%) | 42 (50.6%) | 14 (37.8%) |
| **Pancreatic exocrine status** |  |  |  |
| *Insufficient* | 265 (88.3%) | 77 (92.8%) | 32 (86.5%) |
| *Faecal elastase (U/g)* | 43 [15, 415]<br>(n=73) | 27.5 [15, 386.5]<br>(n=18) | 127 [33, 335.8]<br>(n=12) |
| *Pancreatic Enzyme Replacement Therapy Use* | 252 (84%) | 73 (88%) | 30 (81.1%) |
| *Lipase Dose those on Pancreatic Enzyme Replacement Therapy (IU/kg/day)* | 4187.7 [2632.1, 6162.9] | 4444.5 [3330.1, 6890.1] | 4683.4 [2152.3, 5990.9] |
| **CF related diabetes** | 113 (37.7%) | 36 (43.4%) | 17 (45.9%) |
| **Gastrointestinal history** |  |  |  |
| *Meconium ileus* | 41 (13.7%) | 15 (18.1%) | 7 (19.4%) |
| *Distal intestinal obstruction syndrome* | 62 (20.8%) | 24 (28.9%) | 10 (27%) |
| *Gastroesophageal reflux disease* | 130 (43.3%) | 44 (53%) | 16 (43.2%) |
| *Gastroesophageal reflux disease and on acid suppressant* | 123 (41%) | 41 (49.4%) | 16 (43.2%) |
| **CF related liver disease** | 95 (31.7%) | 33 (39.8%) | 7 (18.9%) |

**Table 2 -.**
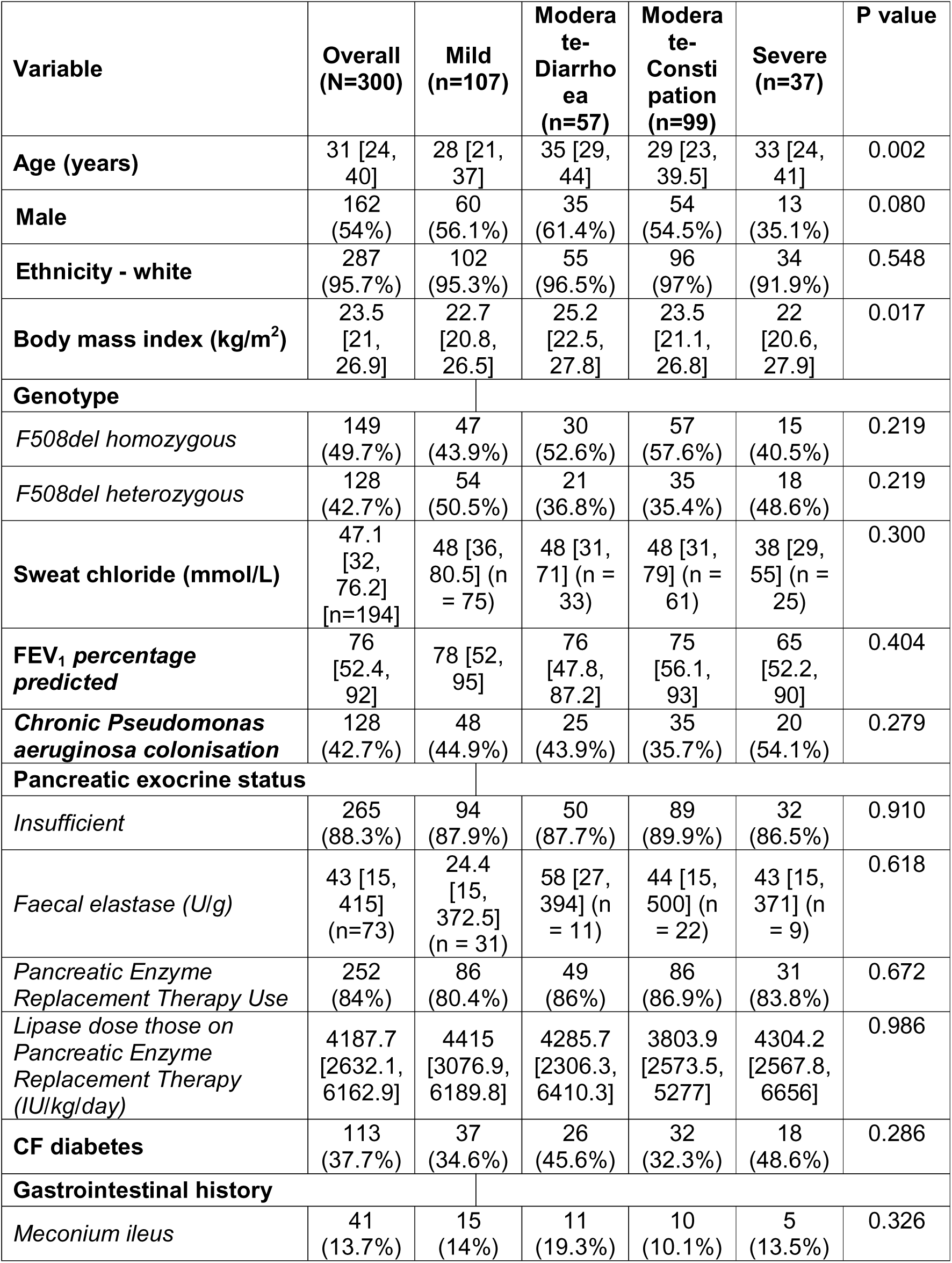
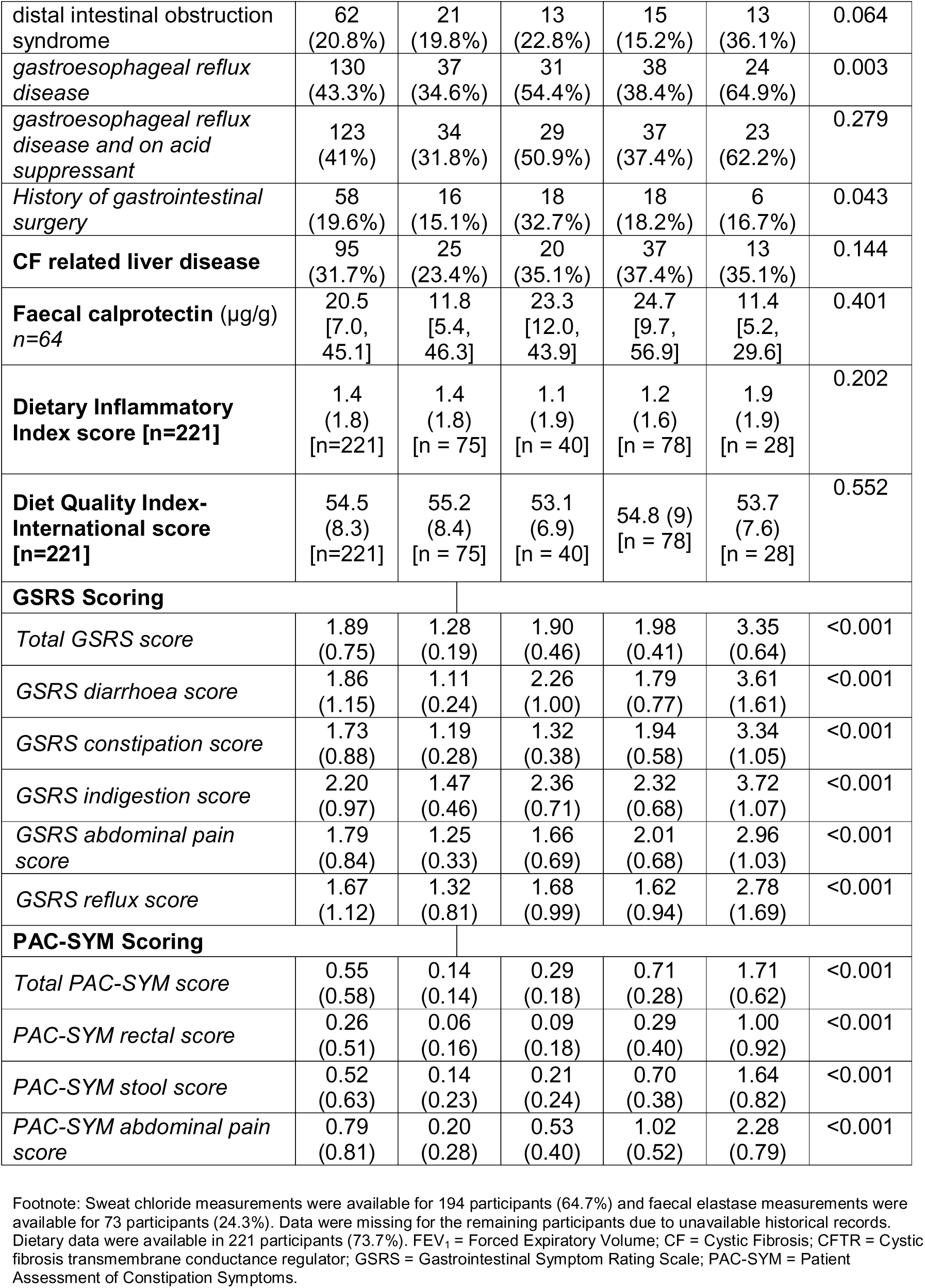
Demographic and clinical characteristics stratified by LCA phenotype. Results are described as mean (SD), median [interquartile range] or number (%)

### Overall cohort symptom scores

Overall, GSRS scores ranged from 1 to 5.27 (maximum score 7), with a mean score of 1.89 (SD 0.75). At least one GI symptom was reported by all 300 participants. Of these, 291 reported at least one symptom specifically on the GSRS or PAC-SYM questionnaires. The most prevalent symptoms across patient-reported outcome measures were bloating (GSRS – 73%; PAC-SYM – 66%), flatulence (65%) and abdominal pain (GSRS – 55%).

### Identification of gastrointestinal symptom phenotypes

#### Model selection

Latent class analysis was performed on responses from 300 participants across 36 symptom indicators, yielding a maximum of 10,800 data points. Missing values were minimal (3%, n=329). A four-class model was selected as optimal as it demonstrated good model fit with balanced high entropy (0.931), indicating minimal classification uncertainty and minimised overfitting (Figure 2a-b, d). All classes met the minimum 10% prevalence criterion (Figure 2c). While models with three classes provided similar results to four-class, the four-class model offered further distinction. Models with five or six classes showed marginal improvements in fit but produced classes with <10% prevalence and suggested more overfitting with reduced clinical utility (Figure 2).

**Figure 2.**
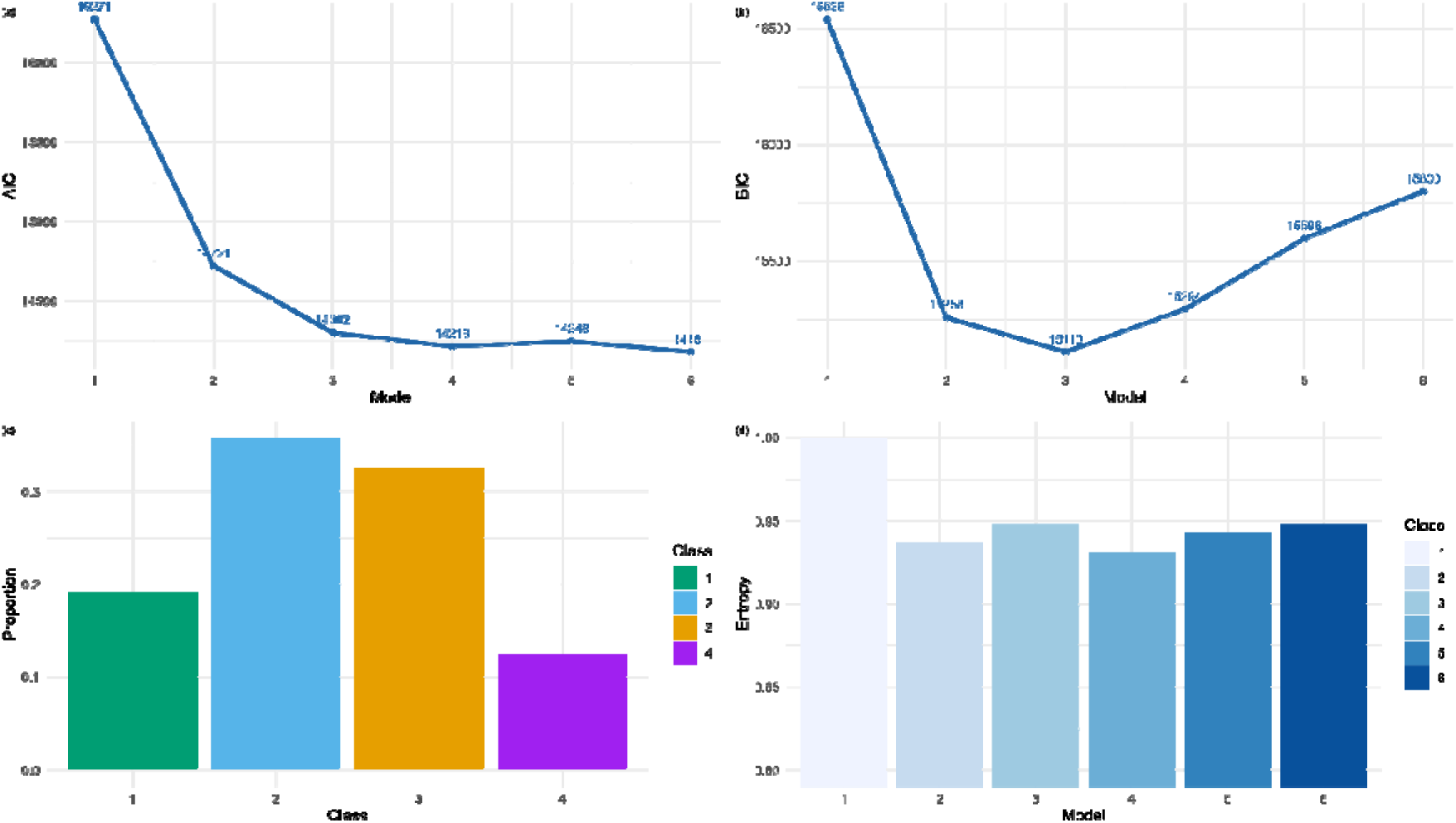
Comparison of model fit across class models and estimated latent class memberships: Line graphs and bar plots comparing (a) Akaike information criteria (AIC), (b) Bayesian information criteria (BIC), and (d) entropy. (c) Bar plot showing the estimated latent class membership proportions

#### Phenotype characteristics

Based on posterior probabilities of class membership (Figure S2-4), four distinct GI symptom phenotypes were identified: mild (Class 2, 36%, n=107); moderate constipation-predominant (Class 3, moderate-C; 33%, n=99); moderate diarrhoea-predominant (Class 1, moderate-D; 19%, n=57) and severe (Class 4, 12%, n=37). The mean posterior probability of class assignment was 0.89 (range 0.76–0.98), indicating robust classification certainty across classes.

The severe phenotype was characterised by a higher probability of scoring 3 (severe) than the other phenotypes across most indicators. In contrast, the mild phenotype showed predominantly mild scores (1) across indicators.

The two moderate phenotypes showed intermediate symptom severity across most indicators but were distinguished by specific symptom patterns. The moderate-C phenotype displayed higher scores for constipation-related symptoms (hard stools, straining, incomplete evacuation, and feeling unable to pass a bowel movement), whereas the moderate-D phenotype was characterised by higher scores for diarrhoea, loose stools and urgency.

The probability trends observed were reflected in the mean GI symptom score of each indicator, presented in Figure 3, emphasising the differences in symptom scores across phenotypes. Further distinction between the two moderate phenotypes was demonstrated and emphasised by significant differences in mean scores across the constipation and diarrhoea related indicators (Figure S5)

**Figure 3.**
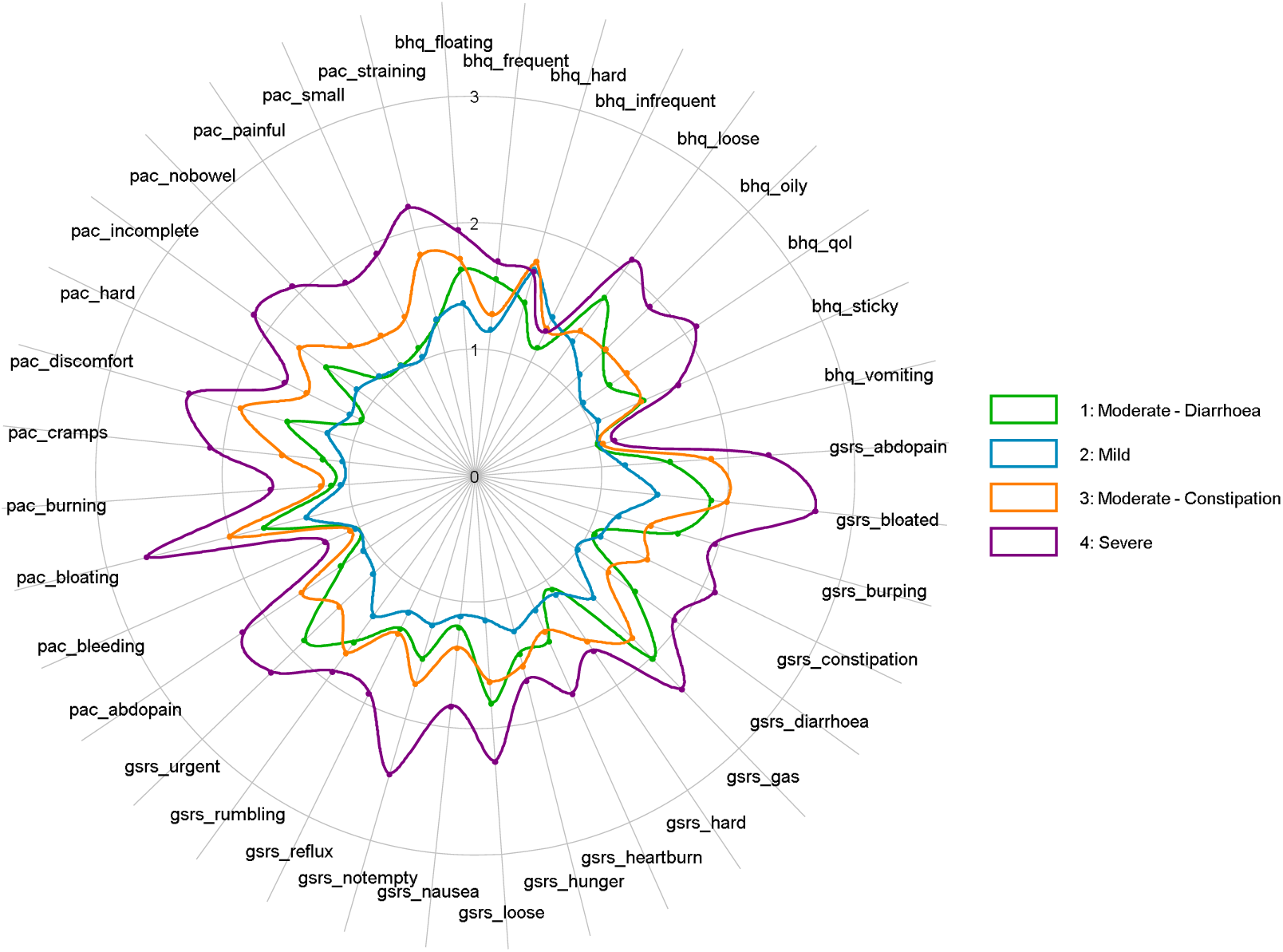
Radar plot comparing mean baseline line scores for adults with CF with mild (blue), diarrhoea-predominant moderate (orange), constipation-predominant moderate (green) and severe (purple) phenotypes

#### Demographic and Clinical Characteristics by Latent Class Phenotype

There were no significant differences between phenotypes in sex or ethnicity (Table 4). Similarly, CFTR genotype distribution (proportion with at least one F508del variant versus other genotypes), and pancreatic insufficiency status did not differ significantly across groups. Sweat chloride concentrations, FEV_₁_ percent predicted and rates of chronic *Pseudomonas aeruginosa* infection were also comparable across phenotypes. Faecal calprotectin levels were similar across phenotypes and were predominantly within normal ranges. Faecal elastase variability was high, and no significant difference was observed between phenotypes. The validated GI symptom questionnaires of GSRS and PAC-SYM overall scores were greater in the severe phenotype, results were more comparable between mild and moderate phenotypes (Table 2).

Dietary recall of at least one day was completed by 222 participants, and one participant’s data was excluded due to data quality issues. The overall mean DQI-I score was 54.5 (SD 8.3) (Table 2). Overall DQI-I scores were similar across groups, the moderate-D phenotype had the lowest mean score 53.1 (SD 6.9) and the mild phenotype had the highest mean score 55.2 (SD 8.4). The mean DII score was 1.4 (SD 1.8), with individual scores ranging from -3.31 (anti-inflammatory) to 4.85 (pro-inflammatory). The moderate-D phenotype had the lowest mean DII score of 1.1 (SD 1.9), whilst the severe phenotype had the highest mean DII score of 1.9 (SD 1.9).

Whilst differences were descriptively apparent (Figure 4, Table 2), no statistical significance was observed.

**Figure 4.**
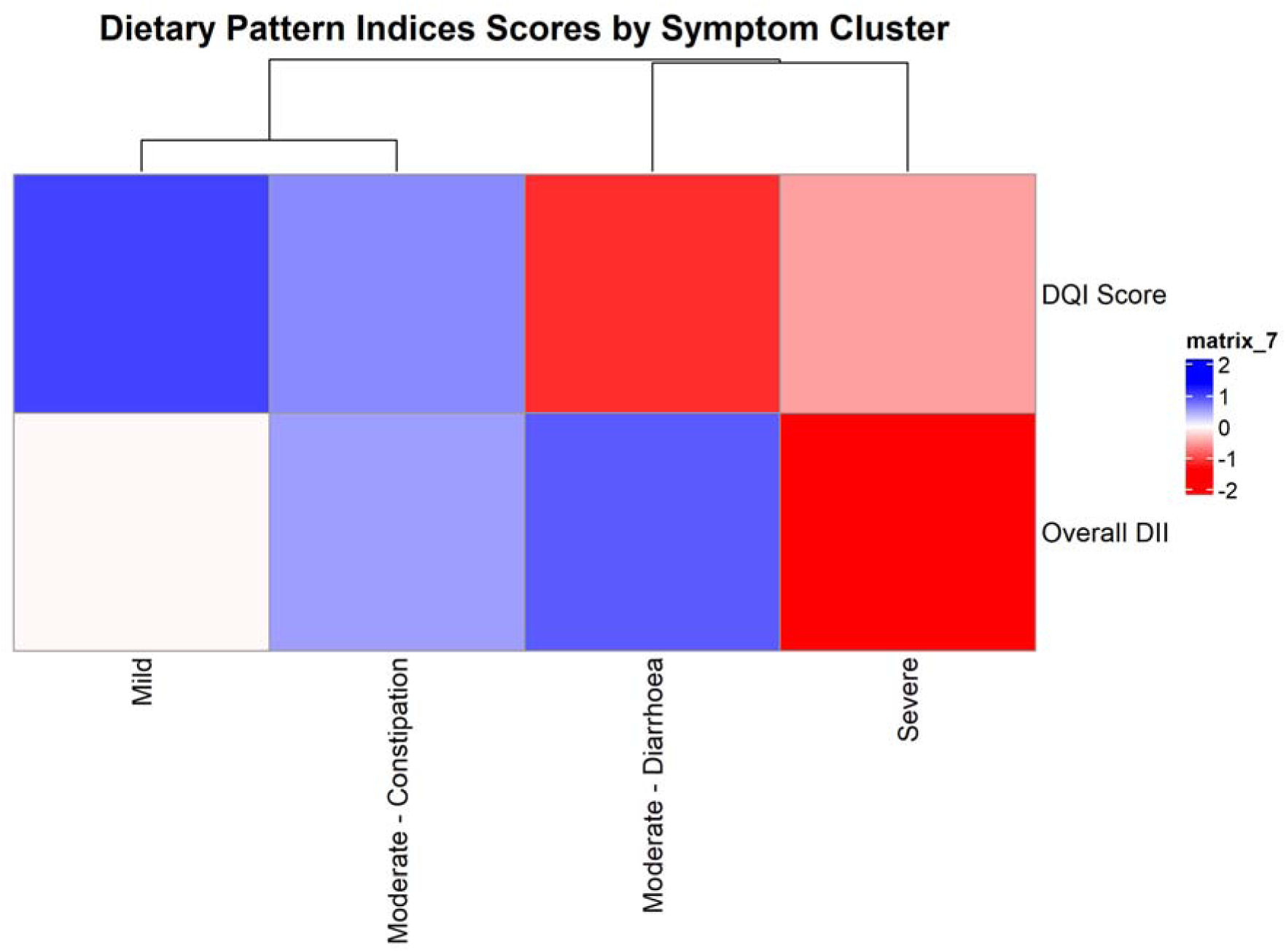
heatmap of mean scores of dietary pattern indices stratified by gastrointestinal phenotype. Dietary inflammatory index score is reversed for interpretation of colours. Red = poorer diet quality measured using DQI-I and higher potential pro-inflammatory

Relative to the mild phenotype, the severe phenotype was associated with gastroesophageal reflux disease (RRR 2.86; 95%CI 1.30-6.31; p=0.009) and distal intestinal obstruction syndrome (RRR 2.46; 95%CI 1.04-5.81; p=0.04) (Figure 5a, supplemental Table S3 c, d). CF-related liver disease was associated with both the moderate-C phenotype (RRR 2.08; 95%CI 1.13-3.81; p=0.018) and the moderate-D phenotype (RRR 2.11; 95%CI 1.03-4.29; p=0.04) (Figure 5a, supplemental table S3 f).

**Figure 5.**
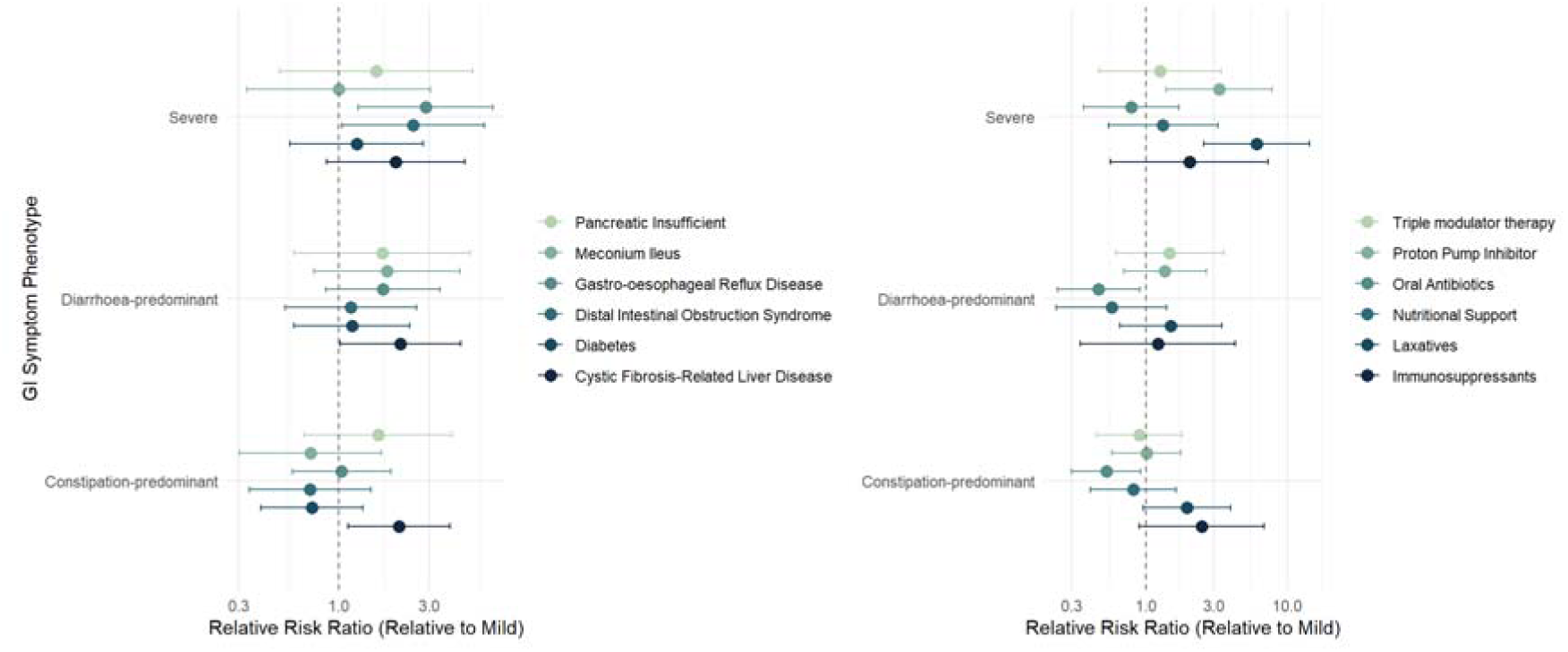
Forest plot showing results of multinomial logistic regression of complications and treatments on GI phenotype, adjusted for age and sex relative to the mild phenotype. Complications: Meconium ileus, Pancreatic insufficiency, gastroesophageal reflux disease, Distal Intestinal Obstruction Syndrome, Diabetes, Cystic Fibrosis-Related Liver Disease. Treatment: Proton Pump Inhibitor, Oral antibiotics, Nutritional support, Laxatives, Immunosuppressants, Triple modulator therapy

Relative to the mild phenotype, the severe phenotype was significantly associated with the use of proton pump inhibitors (RRR 3.29; 95%CI 1.39-7.74; p=0.007) and laxatives (RRR 6.13; 95%CI 2.54-14.84; p<0.001) (Figure 5b, supplemental table S4 b, d). There was a lower likelihood of long-term oral antibiotic use (primarily azithromycin) in both the moderate-C phenotype (RRR 0.53; 95%CI 0.3-0.92; p=0.025) and moderate-D phenotype (RRR 0.46; 95%CI 0.24-0.91; p=0.025), relative to participants in the mild phenotype (Figure 5b, supplemental table S4 c).

## DISCUSSION

We identified and classified four novel and distinct GI phenotypes in adults with CF using LCA, representing the first systematic application of multivariate clustering methods to GI symptoms in this population. The four phenotypes were mild; moderate constipation predominant (moderate-C); moderate diarrhoea-predominant (moderate-D); and severe. Critically, these phenotypes showed differential associations with specific GI complications and medication requirements, suggesting distinct underlying pathophysiology.

Demographic characteristics, CFTR genotype distribution and pancreatic insufficiency status were comparable across phenotypes, indicating that GI symptom heterogeneity is not explained by these traditional CF disease markers. These findings establish that GI symptom presentation in CF is heterogeneous and phenotypically complex, warranting a precision medicine approach to symptom management rather than uniform treatment strategies. Previous research in CF focused on identifying individual GI symptoms in PwCF. Consistent with a recent rapid review looking at GI symptoms and complications in CF (2), this research found that the most reported symptoms overall were bloating, flatulence and abdominal pain. The use of LCA to identify symptom phenotypes has proven valuable in other chronic GI conditions, particularly irritable bowel syndrome and liver disease, where distinct clusters have been associated with different pathophysiological mechanisms and treatment responses (13–15, 35). These studies suggest that traditional symptom classification may be too simplistic, whilst LCA allows further stratification and personalised symptom management. Our findings extend the use of LCA to CF for the first time, demonstrating that multivariate symptom patterns provide more clinically relevant classification than single symptom domains or total symptom scores, which can be limited by unidimensional assessment.

The phenotype specific associations with GI complications suggest underlying mechanistic differences. The association of CF-related liver disease was noted in both moderate phenotypes but not observed to be associated with the severe phenotype, relative to mild. CF-related liver disease is linked with portal hypertension and altered gut-liver-axis signalling, which could manifest as altered bowel habit patterns (36). The absence of this association in the severe phenotype, despite more severe overall symptoms, suggests that the severe phenotype may be driven by different mechanisms. Mechanistic differences in the severe phenotype may relate to underlying motility disorders, supported by association with distal intestinal obstruction syndrome, and to acid hypersecretion, as indicated by the gastroesophageal reflux disease association. There are reported associations of gastroesophageal reflux disease and more severe pulmonary disease (37) suggesting gut-lung axis, here we find a higher prevalence of gastroesophageal reflux disease in the severe phenotype but no significant associations with lung function. With regards to treatments, the severe phenotype was significantly associated with proton pump inhibitor use, relative to the mild phenotype. Whilst proton pump inhibitor use may reflect appropriate treatment of reflux symptoms, long-term acid suppression has been associated with GI symptoms (38), altered intestinal microbiome composition and increased risk of intestinal infections, potentially contributing to ongoing symptom burden (39). The high laxative use in the severe phenotype, for which participants reported high on both constipation and diarrhoea features, further suggests GI dysmotility requiring multimodal symptomatic management. Participants in moderate phenotypes were at lower risk of long-term oral antibiotic use relative to the mild phenotype. Long-term azithromycin has immunomodulatory and prokinetic properties beyond its antimicrobial effects (40).

The PROMISE-GI study (41) proposed that dysbiosis and gut inflammation may compromise intestinal barrier integrity, leading to bacterial translocation and liver dysfunction. In the GRAMPUS-CF cohort, the moderate phenotypes showed both elevated CF-related liver disease prevalence and reduced antibiotic exposure. It is possible that the absence of chronic macrolide therapy may permit microbiome configurations associated with both GI symptoms and liver inflammation. Testing these hypotheses will require integrated analysis of microbiome composition, inflammatory markers and intestinal permeability measures across phenotypes, which are planned analyses within GRAMPUS-CF.

Our findings are particularly relevant given the widespread use of highly effective CFTR modulator therapies. Previous studies have reported conflicting effects of triple CFTR modulator therapy on GI symptoms. The European RECOVER study found improvements in GI symptoms following triple therapy introduction using the CF-specific CFAbd-Score (42), whilst the US-based PROMISE-GI study found statistically significant but clinically modest improvements after 6 months of therapy (36). In our cohort, where 80% of participants were established on triple therapy, all 300 participants reported at least one GI symptom, with substantial proportions experiencing moderate or severe symptom phenotypes. This confirms that GI symptom burden persists in the CFTR modulator era and remains an important treatment priority, as emphasised by recent international patient priority-setting exercises (3, 5, 6).

It has been suggested that combinations of foods and nutrients, as described by dietary patterns, may more effectively demonstrate associations with health conditions. The observation that all phenotypes demonstrated suboptimal diet quality (DQI-I scores <60 (31)) and moderately pro-inflammatory dietary patterns is consistent with previous systematic reviews documenting poor dietary quality (16) in adults with CF despite high energy intake. However, the absence of significant differences in dietary indices between phenotypes was unexpected. This may reflect that dietary patterns were shaped primarily by nutritional recommendations focused on energy density rather than diet quality, resulting in uniformly poor dietary patterns across phenotypes. The severe phenotype exhibited the most pro-inflammatory dietary pattern (DII score 1.9), though this did not reach statistical significance.

Cross-sectional studies of adults with functional constipation have demonstrated that pro-inflammatory diets are associated with altered gut microbiota composition (43), suggesting that dietary factors may play a permissive rather than deterministic role in symptom phenotype development.

### Strengths and limitations

This study has several important strengths and limitations. The cross-sectional nature of the baseline data precludes inference of causal relationships. Longitudinal follow-up will be examined in future publications to assess the stability of gastrointestinal phenotypes over time.

This study represents the first application of LCA to systematically define GI symptom phenotypes in CF, employing robust statistical methodology with high classification certainty. This provides a new framework for investigating the biological mechanisms underlying symptom heterogeneity and for developing precision focused therapeutic strategies. LCA methodology offers probability-based class assignment in modelling categorical ordinal data whilst accommodating measurement similarity (34).

The selection of symptom assessment measures was informed by the systematic review of GI symptoms reported in CF (2), ensuring that key gastrointestinal symptoms highlighted in the literature were comprehensively captured, and could be comparable to other studies(36, 42, 44). This included the BSFS (26) in the questionnaire panel, as well as GSRS (24, 25) and the PAC-SYM (22, 23) questionnaires, which have been successfully applied in CF populations (44–46). Whilst these patient-reported outcome measures are validated and minimally important differences have been previously investigated, self-reporting may introduce bias from subjective perception. Pre-processing steps standardised inconsistent Likert scales and directionality into a simple 3-point score, which minimises impact of differential item functioning at a potential loss of more granular perceptions.

The large sample size was specified to enable reliable phenotype identification in the overall cohort. Subgroup allocation was voluntary and determined by participant preference, which could introduce selection bias. Furthermore, due to restricted sample sizes within sampling subgroups and engagement with additional self-reporting tools, power to identify statistically significant differences in diet and inflammatory markers was limited. Where significant effects were observed with clinical characteristics, caution is needed in interpreting the direction of effect and causal inference. Medication use reflects prescribing patterns and may not fully capture medication adherence or over-the-counter use. Diagnosis of gastroesophageal disease and CF related liver disease was taken from medical records and may not reflect gold standard diagnostic criteria.

The multi-centre design enhances generalisability whilst the nested subgroup structure enables future mechanistic investigation through integrated multi-omics analyses linked at the individual level. The cohort is representative of the contemporary CF population, with high prevalence of CFTR modulator therapy and typical genotype distribution, supporting clinical relevance. However, the GRAMPUS-CF cohort was predominantly white, potentially limiting generalisability to more ethnically diverse populations. Given the phenotypic overlap with irritable bowel syndrome, the absence of psychological assessment constitutes a limitation, as affective disorders may affect symptom perception (14). Future studies are necessary to independently validate the four GI phenotypes in other CF populations, whilst longitudinal follow-up will enable examination of phenotype stability over time.

### Future directions

Integration of microbiome data, faecal inflammatory markers and MRI measures from nested subgroups will enable testing of specific mechanistic hypotheses regarding motility, inflammation and gut-liver-axis dysfunction (47) (supplementary Figure S6). These phenotype definitions can inform targeted intervention studies, including prokinetic therapy for severe phenotype and microbiome-targeted therapies for moderate phenotypes.

We also plan to evaluate phenotype stability. LCA using the same model, will be repeated using GI symptom data collected via 6-month and 12-month follow-up questionnaires which consists of the same questions. Lastly, validation studies will be conducted to validate the LCA model created and examine reproducibility and robustness using a secondary cohort.

In conclusion, this study demonstrates that GI symptom presentation in CF is heterogeneous and phenotypically structured, with distinct clinical associations suggesting different underlying mechanisms. This phenotype-driven approach addresses a critical unmet need and provides a foundation for precision-based GI symptom management strategies.

## Abbreviations

Akaike Information Criterion: AIC
Bayesian Information Criterion: BIC
Bristol Stool Form Scale: BSFS
Bowel habits questions: BHQ
Cystic Fibrosis: CF
Cystic fibrosis transmembrane conductance regulator: CFTR Confidence intervals CI
Dietary Inflammatory Index: DII
Diet Quality Index –: International DQI-I
Forced Expiratory Volume in one second: FEV₁
Forced Vital Capacity: FVC
Gastrointestinal: GI
Gut Research Advancing a Mechanistic and Personalised Understanding of Symptoms in Cystic Fibrosis: GRAMPUS-CF
Gastrointestinal Symptom Rating Scale: GSRS
Interquartile range: IQR
Latent Class Analysis: LCA
Magnetic resonance imaging: MRI
Patient Assessment of Constipation Symptoms: PAC-SYM
People living with cystic fibrosis: PwCF
Relative Risk Ratio: RRR

### Disclosures

This author discloses the following: DP speaker/board honoraria from Vertex RS Research Grants from Sanofi and Nestle, Consultant to Enterobiotix. Alan Smyth reports support for this study from UK CF Trust Strategic Research Centre Grant (SRC023); patent on Alkyl quinolones as biomarkers of Pseudomonas aeruginosa infection and uses thereof (Pub-Chem Patent Summary US-2016131648-A1 https://pubchem.ncbi.nlm.nih.gov/patent/US-2016131648-A1); speaker honoraria from Vertex Pharmaceuticals (paid to institution); and participation on the Data Safety Monitoring Board and US Cystic Fibrosis Foundation (2019-present).

No relevant conflicts of interest exist for the following authors: JH, DS, HS, CJ, AR, RM, AP, TM, LM, HB, DD, CvdG,

### Authors

**Holaman, J**. (Data curation: Lead; Formal analysis: Lead; Investigation: Lead; Visualization: Lead; Writing – original draft: Lead)

**Sills, D,** (Data curation: Lead; Formal analysis: Lead; Investigation: Lead; visualization: Lead; writing – Original draft: Lead; Project administration: Lead)

Saumtally, H (Data curation: Lead; Investigation: Lead; Writing – review & editing: Equal; Project administration: Lead)

Johnson, C, (Writing – review & editing: Supporting; Investigation: Lead) Recto, A, (Writing – review & editing: Supporting)

Marsh, R, (Writing – review & editing: Supporting)

Prayle, A, (Supervision: Lead; Writing – review & editing: Supporting)

Monaghan, T, (Conceptualisation: Lead; Supervision: Lead; Funding acquisition: Supporting; Writing – review & editing: Equal)

Marciani, L, (Conceptualisation: Lead; Supervision: Lead; Funding acquisition: Supporting; Writing – review & editing: Equal)

Spiller, R, (Conceptualisation: Lead; Supervision: Lead; Funding acquisition: Supporting; Writing – review & editing: Equal)

Barr, H.L, (Supervision: Lead; funding acquisition: Supporting; Writing – review & editing: Equal)

Downey, D. G., (Investigation: Lead; Writing – review & editing: Equal; Project administration: Lead)

van der Gast, C, (Conceptualisation: Lead; Supervision: Lead; Funding acquisition: Supporting; Writing – review & editing: Equal)

Peckham, D (Conceptualisation: Lead; Supervision: Lead; Funding acquisition: Supporting; Writing – review & editing: Equal)

Stewart, I (Conceptualisation: Lead; Data curation: Lead; Formal analysis: Lead; Supervision: Lead; Visualization: Lead; Methodology: Lead; Writing – original draft: Supporting)

Smyth, A (Conceptualisation: Lead; Supervision: Lead; Methodology: Lead; Writing – original draft: Supporting; Project administration: Lead)

**"Author names in bold designate shared co-first authorship" at the end of the references section.**

### Data Transparency Statement

Requests should be addressed to the chief investigator via the study email address. Requests will be assessed on a case-by-case basis.

Applications should state the research question being addressed and include a link to the researcher’s published protocol. This will be reviewed by the research team and a final decision to share data will be the responsibility of the chief investigator. Data sharing is specifically mentioned in the participant information sheet and consent for this has been obtained.

Applications will be considered from the time that our own data analysis is complete (expected to be 31/12/26), for a maximum of 7 years after study completion.

