## Supplementary document for "Distinct Gastrointestinal Symptom Phenotypes in Adults with Cystic Fibrosis are linked to complications and medication use"

**Supplementary materials:**

**Methods**

Adult clinic population

Leeds n=386

Nottingham n=210

Belfast n=279

GROUP A

Completed questionnaires

n=300

Group B

Stool & Blood samples for Microbiome & inflammatory mediators

n=83

Group C

Detailed inflammatory phenotyping & MRI

n=37

Figure S1 GRAMPUS-CF study design: Overview of the nested study structure, including groups and subgroups.

Table S1 Description of GRAMPUS questionnaire panel.

| Patient-reported outcome measures/ Questionnaire | Description | Domains | Scale |
| --- | --- | --- | --- |
| PAC-SYM (22, 23) | Constipation specific focusing on characteristics of constipation (12 items) | Abdominal, rectal and stool | 5-point Likert scale |
| GSRS (24, 25) | Gastrointestinal symptom assessment tool used to evaluate common symptoms of gastrointestinal disorders (15 items) | Abdominal pain and reflux, diarrhoea, indigestion, and constipation syndromes | 7-point Likert scale |
| ‘GRAMPUS seven’ Bowel habits questions | Bowel-specific focusing on characteristics of bowel habits (7 items) including the Bristol Stool Form Chart (26) | Bowel movement | 3-point Likert scale  Number of bowel movements per day was rated |

**GRAMPUS Seven questions:**

The ‘GRAMPUS seven’ questions used a combination ‘times per day…’ and a 3-point frequency-based Likert-scale (often, occasionally and never). The questions were supplemented with the use of the Bristol Stool Form Scale (BSFS) (26) . The seven questions asked participants:

1. On average, how many times per day did you have a bowel movement, in the last week? (answers 0-6 or more)
2. In the last week were your stools (poo) – list of Bristol Stool Frequency Chart stool types (answers often, occasionally, never)
3. In the last week has your stool (poo) been pale and sticky? (answers often, occasionally, never)
4. In the last week did you have a stool (poo) which floats in the toilet? (answers often, occasionally, never)
5. In the last week have you seen oil floating on the water in the toilet after you have passed stool (poo)? (answers often, occasionally, never)
6. In the last week have you had any episodes of vomiting? (answers often, occasionally, never)
7. In the last week, have any gut symptoms affected or interfered with your life in general? (answers often, occasionally, never)

**LCA score recoding**

A 3-point Likert scale was chosen because it was the maximum scale possible, as the GRAMPUS-CF bowel habits questionnaire has a 3-point Likert scale.

For each of the patient-reported outcome measures, no discomfort at all and absence of symptoms were scored as mild (1) and responses including “severe” were scored as severe (3). All others were scored as moderate (2).

Unidirectional

GSRS

- No discomfort at all	1
- Minor discomfort 	2
- Mild discomfort 	2
- Moderate discomfort 	2
- Moderately severe discomfort 	3
- Severe discomfort 	3
- Very severe discomfort 	3

PAC-SYM

- Absence of symptoms	1
- Mild 	2
- Moderate 	2
- Severe 	3
- Very severe	3

GRAMPUS-CF bowel habits questionnaire

- Never	1
- Occasionally 	2
- Often 	3

Bidirectional

Questions 1 and 2 of the GRAMPUS-CF bowel habits questions were bidirectional. For the LCA, two variables were created for each question: frequent and infrequent, and hard and loose for questions 1 and 2, respectively.

Question 1

On average, how many times per day did you have a bowel movement, in the last week?

Infrequent

1 – 2, 3, 4, 5, 6

2 – 1

3 – 0

Frequent

1 – 0, 1

2 – 2, 3

3 – 4, 5, 6*

*6 refers to 6 or more as stated in the questionnaire

Question 2 – Hard and loose

Table S2 Recoding rules for items of Bristol Stool Chart

| 2. In the **last week** were your stools (poo) (If you have had more than one type of stool (poo), please circle all that apply) | | Circle one word on each line A-G  All other options 1 | | |
| --- | --- | --- | --- | --- |
| A | 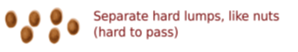 | Often  Hard 3 | Occasionally  Hard 2 | Never  Hard 1 |
| B | 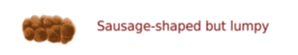 | Often  Hard 2 | Occasionally | Never  Hard 1 |
| C | 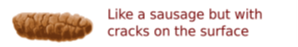 | Often | Occasionally | Never  Loose 2 |
| D | 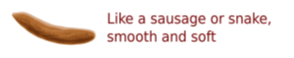 | Often | Occasionally | Never  Hard 2  Loose 2 |
| E | 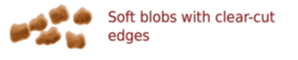 | Often | Occasionally | Never  Hard 2 |
| F | 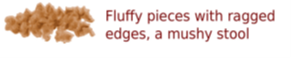 | Often  Loose 2 | Occasionally | Never  Loose 1 |
| G | 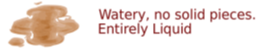 | Often  Loose 3 | Occasionally  Loose 2 | Never  Loose 1 |

**Diet quality checks and cleaning:**

Each food item, alongside any additional prompted items was recorded. The output includes the participant’s original search term, confirmed food description, associated nutrient table name and food group, serving size (in grams or millilitres), amount of leftovers and an indication of whether the reported portion size was considered reasonable. When Intake24 could not identify a food item, participants could manually enter a food description, portion size, and leftovers. Based on the food entries and quantities consumed, nutrient values were calculated for each item in the file.

Some food entries were recorded as composite meals and required disaggregation into individual components. For example, an entry such as “salt and pepper chicken with egg fried rice” was split into separate items for salt and pepper chicken and egg fried rice to allow more accurate nutrient profiling. Where food items were missing, efforts were made to match them with existing entries in the raw data. If no suitable match was available, a supplemental CSV file from Intake24 was used to look up and extract appropriate nutrient values. These matched values were then imported into the main dataset to ensure completeness.

In cases where foods could not be identified via Intake24, additional nutrient information was sourced directly from food manufacturers or commercial providers. These entries typically included a reduced set of nutrient data often limited to energy, carbohydrate, sugar, fat, saturated fat, protein and fibre. Where information remained sparse, estimates were made based on the participant’s previous dietary entries and standard knowledge of commonly consumed foods.

Portion sizes were assessed using a combination of the participant’s overall dietary intake and guidance from the Food Standards Agency on typical portion sizes across different food groups. This step ensured consistency and plausibility in reported intakes.

A series of monitoring checks were applied to highlight data issues and assess overall quality. These included identifying recalls with fewer than ten items, recalls completed in under two minutes and energy intakes falling below 400 kcal or exceeding 4000 kcal. Instances of missing foods were also logged for manual investigation.

To detect implausibly high intakes, box plots were used to identify outliers, applying the standard criterion of three times the interquartile range. Recalls flagged as outliers were then reviewed to determine whether the elevated energy intake could be explained by a specific food item. Decisions to retain, adjust or exclude such records were informed by knowledge of reasonable daily energy intakes based age on and sex, as well as the plausibility of reported entries. An additional check focused on implausible portion sizes. These outliers were manually reviewed and depending on contextual plausibility, were either accepted, amended or excluded from the analysis after confirmation with a second Dietitian.

**Results**

***Item-response probabilities***

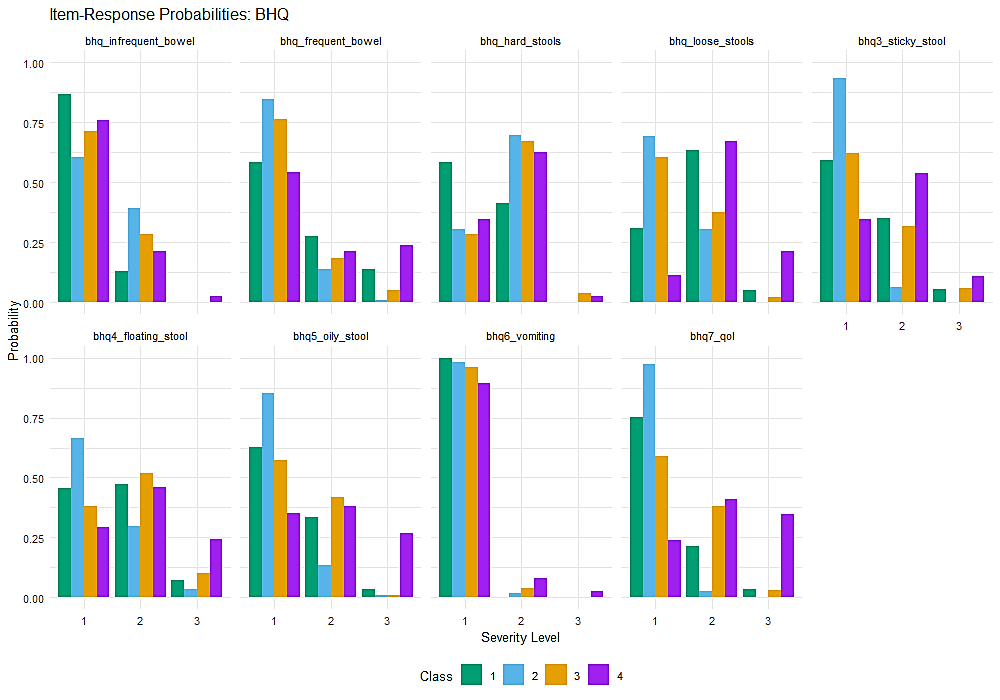

f

g

h

i

e

d

c

b

a

Figure S2 Item-response probabilities derived from responses (1 – 3) to 7 bowel habits questions (BHQ) and 9 indicators. Item-response probabilities derived from responses (1 – 3) to 7 bowel habits questions (BHQ) and 9 indicators. Class 1 = Moderate-constipation phenotype, Class 2 =Mild phenotype, Class 3 = Moderate-diarrhoea phenotype and Class 4 = Severe phenotype.

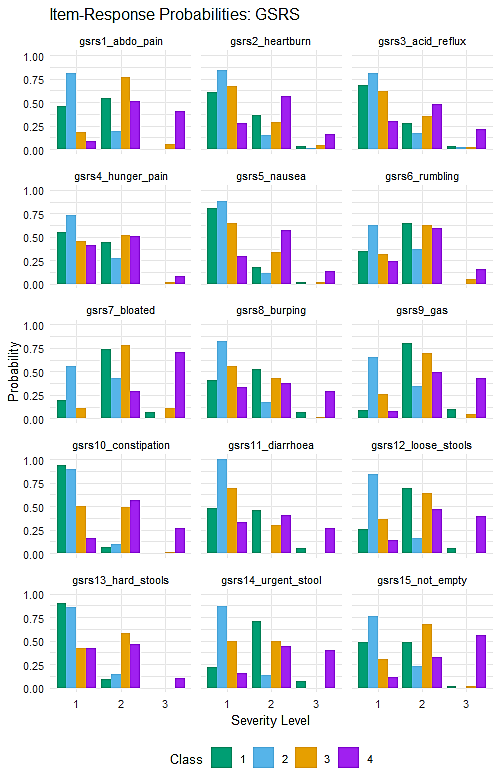

o

n

m

l

k

j

i

h

g

f

e

d

c

b

a

Figure S3 Item-response probabilities derived from responses to GSRS items. Class 1 = Moderate-constipation phenotype, Class 2 =Mild phenotype, Class 3 = Moderate-diarrhoea phenotype and Class 4 = Severe phenotype.

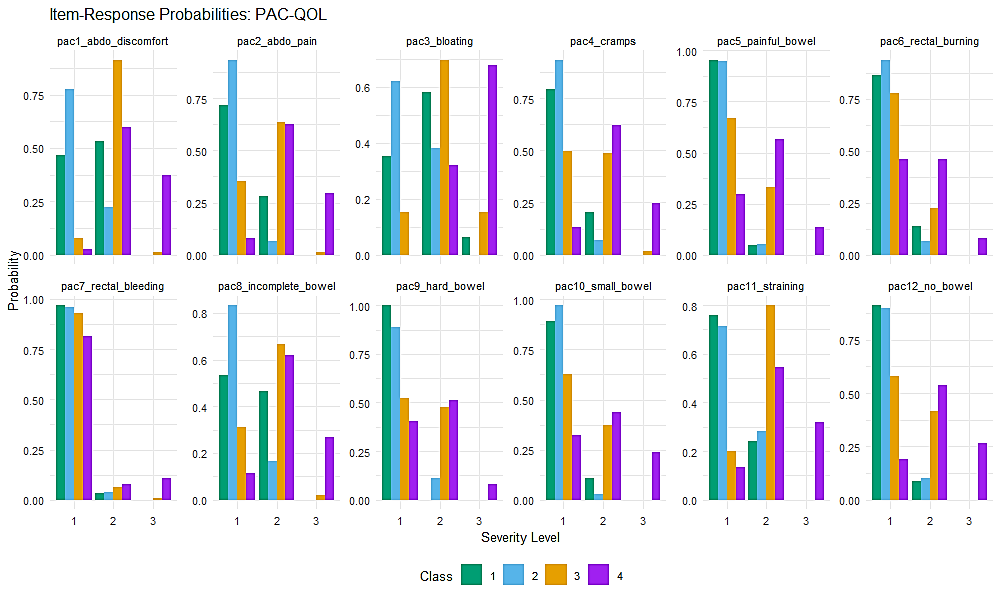

Figure S4 Item-response probabilities derived from responses to PAC-SYM items. Class 1 = Moderate-constipation phenotype, Class 2 =Mild phenotype, Class 3 = Moderate-diarrhoea phenotype and Class 4 = Severe phenotype.

***Mean baseline scores for moderate phenotypes***

a

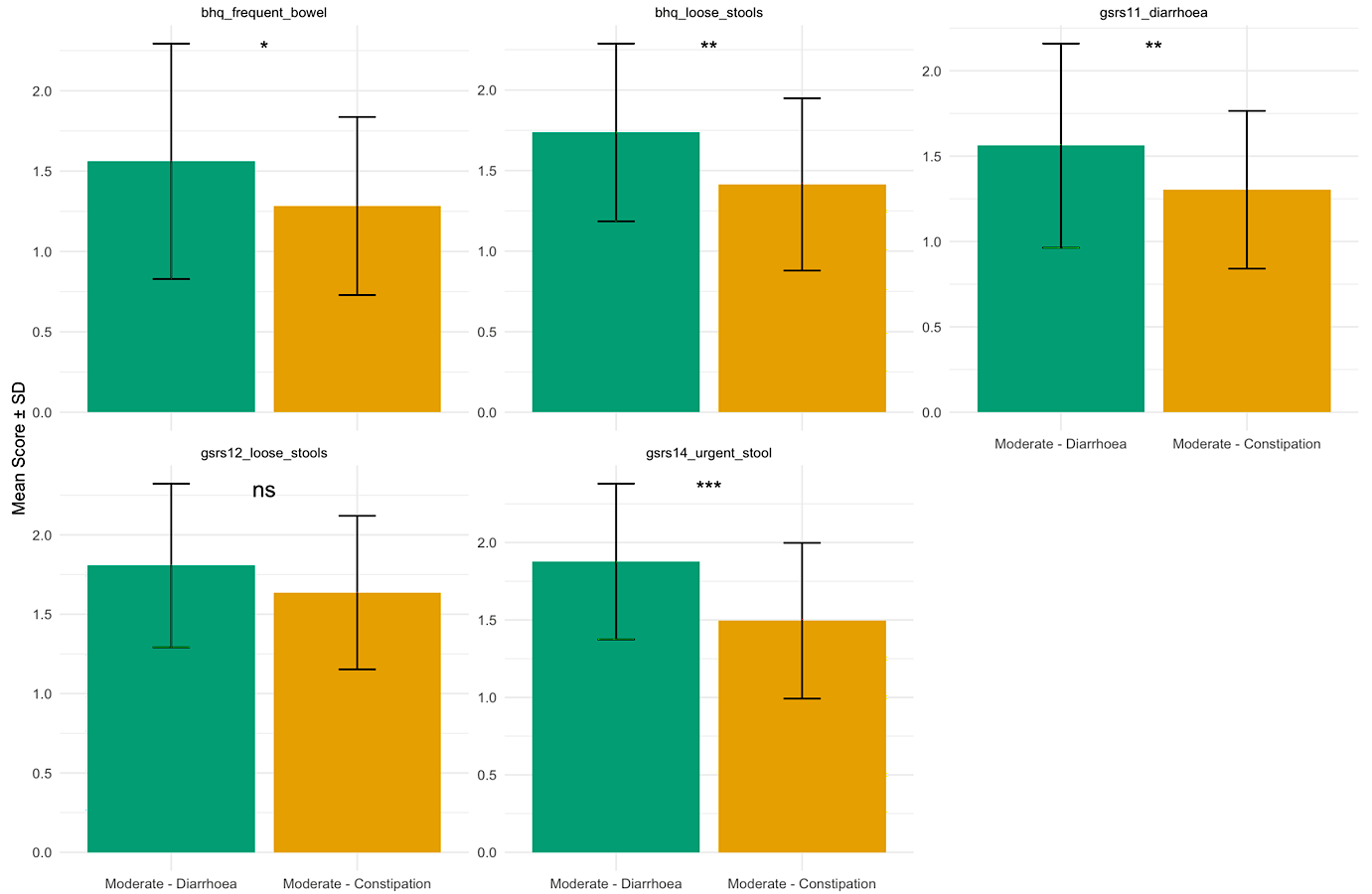

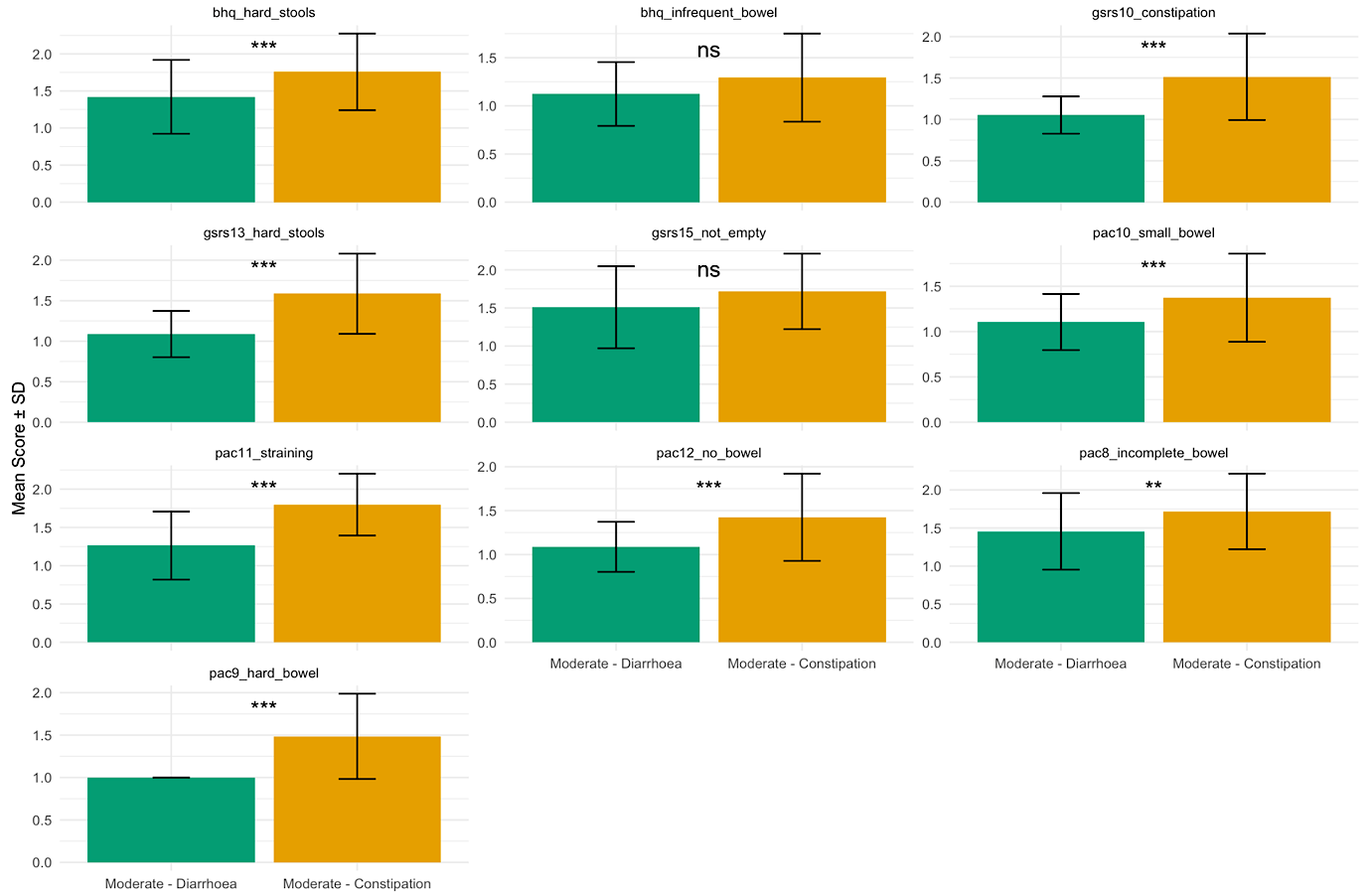

b

Figure S5 Bar plots of mean baseline scores per Moderate phenotype. Moderate phenotypes were distinguished by significant differences (* p < .05, ** p < .01, *** p < .001) in mean scores per diarrhoea (a) and constipation (b) related items.

**Figure S6** Mechanistic diagram of pathophysiology of gut symptoms and complications in CF. We propose these arise through a combination of viscid small bowel content, dysbiosis, inflammation, stasis and dysmotility (47).

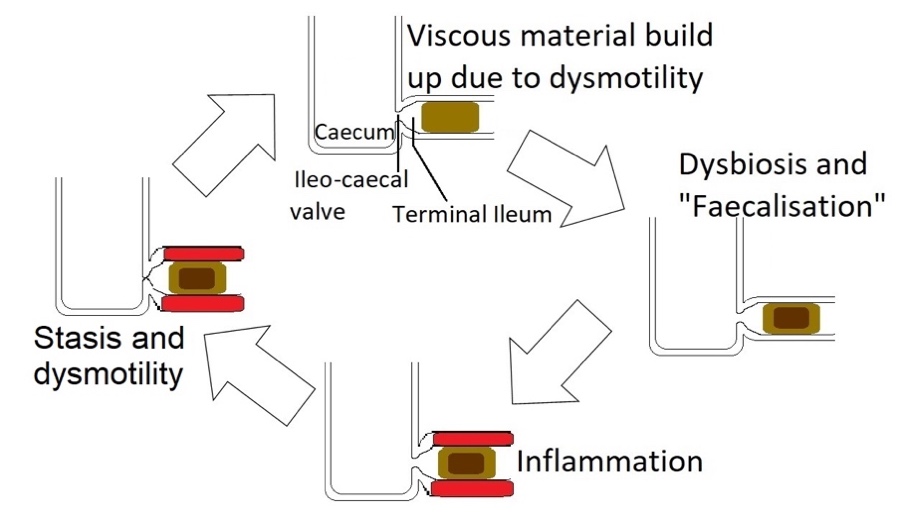

**Relative Risk Ratios – Complications and Treatments**

Table S3 Relative Risk Ratios of cystic fibrosis-related complications relative to the Mild phenotype: (a) Pancreatic Insufficient, (b) Meconium Ileus, (c) Gastro-oesophageal Reflux Disease, (d) Distal Intestinal Obstruction Syndrome, (e) Diabetes, (f) Cystic Fibrosis-Related Liver Disease

| **Pancreatic Insufficient** | | | | | | |
| --- | --- | --- | --- | --- | --- | --- |
| **Phenotype** | **RRR** | **Std.error** | **Statistic** | **P value** | **Low** | **High** |
| Diarrhoea-predominant | 1.69 | 0.54 | 0.97 | 0.33 | 0.58 | 4.91 |
| Constipation-predominant | 1.61 | 0.46 | 1.04 | 0.30 | 0.66 | 3.94 |
| Severe | 1.57 | 0.60 | 0.76 | 0.45 | 0.49 | 5.05 |

| **Meconium Ileus** | | | | | | |
| --- | --- | --- | --- | --- | --- | --- |
| **Phenotype** | **RRR** | **Std.error** | **Statistic** | **P value** | **Low** | **High** |
| Diarrhoea-predominant | 1.79 | 0.45 | 1.30 | 0.19 | 0.74 | 4.32 |
| Constipation-predominant | 0.71 | 0.44 | -0.78 | 0.44 | 0.30 | 1.68 |
| Severe | 1.00 | 0.57 | 0.00 | 1.00 | 0.33 | 3.05 |

| **Gastro-oesophageal Reflux Disease** | | | | | | |
| --- | --- | --- | --- | --- | --- | --- |
| **Phenotype** | **RRR** | **Std.error** | **Statistic** | **P value** | **Low** | **High** |
| Diarrhoea-predominant | 1.71 | 0.35 | 1.52 | 0.13 | 0.86 | 3.40 |
| Constipation-predominant | 1.04 | 0.30 | 0.12 | 0.91 | 0.57 | 1.88 |
| Severe | 2.86 | 0.41 | 2.54 | 0.01 | 1.27 | 6.45 |

| **Distal Intestinal Obstruction Syndrome** | | | | | | |
| --- | --- | --- | --- | --- | --- | --- |
| **Phenotype** | **RRR** | **Std.error** | **Statistic** | **P value** | **Low** | **High** |
| Diarrhoea-predominant | 1.16 | 0.41 | 0.36 | 0.72 | 0.52 | 2.57 |
| Constipation-predominant | 0.71 | 0.37 | -0.93 | 0.35 | 0.34 | 1.47 |
| Severe | 2.46 | 0.44 | 2.07 | 0.04 | 1.05 | 5.78 |

| **Diabetes** | | | | | | |
| --- | --- | --- | --- | --- | --- | --- |
| **Phenotype** | **RRR** | **Std.error** | **Statistic** | **P value** | **Low** | **High** |
| Diarrhoea-predominant | 1.17 | 0.36 | 0.45 | 0.66 | 0.58 | 2.36 |
| Constipation-predominant | 0.72 | 0.31 | -1.03 | 0.30 | 0.39 | 1.34 |
| Severe | 1.24 | 0.41 | 0.52 | 0.60 | 0.55 | 2.79 |

| **Cystic Fibrosis-Related Liver Disease** | | | | | | |
| --- | --- | --- | --- | --- | --- | --- |
| **Phenotype** | **RRR** | **Std.error** | **Statistic** | **P value** | **Low** | **High** |
| Diarrhoea-predominant | 2.11 | 0.37 | 2.01 | 0.05 | 1.02 | 4.36 |
| Constipation-predominant | 2.08 | 0.31 | 2.34 | 0.02 | 1.13 | 3.83 |
| Severe | 1.99 | 0.43 | 1.62 | 0.11 | 0.86 | 4.58 |

Table S4 Relative Risk Ratios of cystic fibrosis-related complications relative to the Mild phenotype: (a) Triple Modulator Therapy, (b) Proton Pump Inhibitor, (c) Oral Antibiotics, (d) Laxatives, (e) Nutritional Support, (f) Immunosuppressants

| **Triple modulator therapy (Kaftrio)** | | | | | | |
| --- | --- | --- | --- | --- | --- | --- |
| **Phenotype** | **RRR** | **Std.error** | **Statistic** | **P value** | **Low** | **High** |
| Diarrhoea-predominant | 1.47 | 0.45 | 0.86 | 0.39 | 0.61 | 3.54 |
| Constipation-predominant | 0.90 | 0.36 | -0.30 | 0.76 | 0.45 | 1.80 |
| Severe | 1.26 | 0.51 | 0.46 | 0.65 | 0.47 | 3.39 |

| **Proton Pump Inhibitor** | | | | | | |
| --- | --- | --- | --- | --- | --- | --- |
| **Phenotype** | **RRR** | **Std.error** | **Statistic** | **P value** | **Low** | **High** |
| Diarrhoea-predominant | 1.37 | 0.34 | 0.91 | 0.36 | 0.70 | 2.68 |
| Constipation-predominant | 1.01 | 0.29 | 0.03 | 0.97 | 0.58 | 1.77 |
| Severe | 3.29 | 0.44 | 2.71 | 0.01 | 1.39 | 7.77 |

| **Oral Antibiotics** | | | | | | |
| --- | --- | --- | --- | --- | --- | --- |
| **Phenotype** | **RRR** | **Std.error** | **Statistic** | **P value** | **Low** | **High** |
| Diarrhoea-predominant | 0.47 | 0.34 | -2.24 | 0.03 | 0.24 | 0.91 |
| Constipation-predominant | 0.53 | 0.29 | -2.26 | 0.02 | 0.30 | 0.92 |
| Severe | 0.79 | 0.40 | -0.60 | 0.55 | 0.36 | 1.71 |

| **Laxatives** | | | | | | |
| --- | --- | --- | --- | --- | --- | --- |
| **Phenotype** | **RRR** | **Std.error** | **Statistic** | **P value** | **Low** | **High** |
| Diarrhoea-predominant | 1.50 | 0.42 | 0.96 | 0.34 | 0.66 | 3.44 |
| Constipation-predominant | 1.96 | 0.36 | 1.86 | 0.06 | 0.96 | 3.97 |
| Severe | 6.05 | 0.44 | 4.09 | 0.00 | 2.55 | 14.35 |

| **Nutritional Support** | | | | | | |
| --- | --- | --- | --- | --- | --- | --- |
| **Phenotype** | **RRR** | **Std.error** | **Statistic** | **P value** | **Low** | **High** |
| Diarrhoea-predominant | 0.57 | 0.46 | -1.22 | 0.22 | 0.23 | 1.41 |
| Constipation-predominant | 0.82 | 0.36 | -0.57 | 0.57 | 0.41 | 1.65 |
| Severe | 1.32 | 0.45 | 0.61 | 0.54 | 0.54 | 3.21 |

| **Immunosuppressants** | | | | | | |
| --- | --- | --- | --- | --- | --- | --- |
| **Phenotype** | **RRR** | **Std.error** | **Statistic** | **P value** | **Low** | **High** |
| Diarrhoea-predominant | 1.21 | 0.64 | 0.30 | 0.76 | 0.34 | 4.28 |
| Constipation-predominant | 2.48 | 0.52 | 1.76 | 0.08 | 0.90 | 6.84 |
| Severe | 2.03 | 0.65 | 1.08 | 0.28 | 0.56 | 7.30 |
